# Antibody Responses to SARS-CoV-2 Vaccine in Nursing Home Residents Support a Bi-Annual Update Schedule

**DOI:** 10.1101/2025.01.09.25320262

**Authors:** Alexandra N. Paxitzis, Oladayo A. Oyebanji, Olajide J. Olagunju, Debbie Keresztesy, Mike Payne, Vaishnavi Ragavapuram, Nicholas Sundheimer, Ellen See, Dennis Wilk, Yi Cao, Yasin Abul, Clare Nugent, Evan Dickerson, Tiffany Wallace, Laurel Holland, Aman Nanda, Walther M. Pfeifer, Alejandro B. Balazs, Christopher L. King, Stefan Gravenstein, David H. Canaday, Brigid M. Wilson, Jürgen Bosch

## Abstract

**Background:** The COVID-19 pandemic has greatly affected nursing home residents (NHRs), a vulnerable group with high rates of illness and death. While vaccination is essential for reducing infections and severe outcomes in the short term, it is important to understand how long antibody levels and neutralizing activity last. This understanding will help us create effective public health strategies for the long term. According to current CDC guidelines, individuals over the age of 65 should receive a booster dose six months after their previous vaccination.

**Methods:** This observational retrospective cohort study analyzed post-vaccination serum from samples with up to 400 days of follow-up from 697 NHRs and 127 healthcare workers (HCWs) across Northeast Ohio and Rhode Island. Analyses were conducted to model decay rates of both neutralizing and binding antibody titers and the impact of previous exposures to SARS-CoV-2 on these decay rates.

**Results:** Results indicate that NHRs show Wuhan and Omicron BA.4/5 neutralizing and binding antibody titers diminish significantly from 2 weeks to 12 months post-vaccination. NHRs with prior infection show higher peak antibody titers and slower decay than those naive to infection. Antibody levels after vaccination for infection-naive NHR residents lagged HCW and NHR with prior infection, but then decayed at a similar rate.

**Conclusion:** The immunologic findings in this cohort of NHR are in line with the existing real-world clinical effectiveness data in older individuals and support the CDC recommendation of a bi-annual vaccination to reduce severe COVID-19 outcomes in persons age 65 and older.

## Introduction

The COVID-19 pandemic, caused by the novel beta-coronavirus SARS-CoV-2, has challenged global public health systems since December 2019 with significant and ongoing morbidity and mortality [1]. Older adults, especially those in nursing homes, more often experience severe outcomes, including hospitalization and death, due to age-related immune decline [2], the acquisition of multiple morbidities, medical complexities, and disabilities. Living in the close quarters of a nursing home elevates the risk of viral transmission [3, 4]. Consequently, nursing home residents and staff early in the pandemic comprised over 23% of all COVID-19 related deaths in the US [5].

A better understanding of the SARS-CoV-2 infection and vaccination immune response dynamics will critically inform the optimization of clinical management strategies and the design of vaccination campaigns for this population. The initial immune responses post-vaccination and infection do not reliably predict their durability. Previous findings from our ongoing longitudinal cohort study indicate that NHRs naive to infection have significantly lower anti-Spike and neutralizing antibody titers at both 2 weeks and 6 months after the initial vaccine series than those with prior infection [6, 7]. Seventy percent of infection-naive NHRs reached the pseudovirus neutralization assay lower limit of detection (LLD) by 6 months after vaccination, whereas only 35% with prior infection reached the LLD at the same time point [7]. Neutralizing antibodies deserve consideration when discussing antibody decay and vaccination schedule, as the Omicron variant escapes vaccine-elicited neutralization in both those naive to infection and those with prior infection after the initial vaccine series [8, 9]. We previously reported that in NHRs, the bivalent (BV) vaccination boost that targets the ancestral Wuhan strain and the Omicron variants BA.4/5, increases neutralizing and binding antibody titers which then begin to decline [10]. Between 2022 and 2023 only one annual vaccination was recommended for non-immunocompromised adults under 65. The FDA first authorized the administration of the BV booster beginning in September 2022, the ACIP recommended an additional bivalent booster for adults over 65 beginning in April 2023 [11]. The first monovalent booster using the XBB.1.5 variant was approved and authorized in September 2023 [12]. Despite these recommendations, by February 2024, only 40.5% of nursing home residents had received a dose of this updated XBB1.5 COVID-19 vaccine [13]. The CDC’s National Healthcare Safety Network notes as of Dec 15, 2024 that 63% of NHRs and 93% of NH staff are not up to date with the current vaccination schedule supporting that reduced uptake of the most recent vaccine is an ongoing issue [14].

Here we report on a serologic assessment of antibody decay following serial SARS-CoV-2 vaccination with and without interval documented infection or serologically inferred infection. We discuss the clinical implications for NHRs of antibody kinetics, including the relevance for vaccine schedules, and identify priorities for future research aimed at informing targeted approaches to better protect NHRs from COVID- 19.

## Materials & Methods

We obtained informed consent from subjects and/or their legally authorized representatives in accordance with the ethical standards set forth by the WCG Institutional Review Board for Case Western Reserve University and Rhode Island Lifespan Hospitals (Study #1316159). We complied with the ethical guidelines for research involving human participants. Funding was provided by grant [CDC] 200-2016-91773, U01 CA260539/CA/NCI NIH/United States. Subject samples were from the interval of 19 January 2021 to 27 October 2023.

We quantified anti-Spike IgG binding antibody titers using a bead-multiplex immunoassay for SARS-CoV- 2 WT, Omicron BA.1, BA.4/5 and XBB1.5 variants [8]. Spike protein from WT, BA.1, BA.5, and XBB1.5 variants were conjugated to Luminex microbeads and a Magpix system was used to analyze antibody fluorescent intensity. Mean fluorescence intensity (MFI) was converted to binding antibody units (BAU/mL) for anti-Wuhan spike and arbitrary units (AU/mL) for BA.1, BA.4/5 and XBB1.5 anti-Spike antibodies.

A SARS-CoV-2 pseudovirus neutralization assay was conducted to assess neutralizing antibody activity [7, 8]. Lentiviral particles were pseudotyped with spike protein from WT and Omicron BA.4/5 variants of SARS-CoV-2, and transfected to HEK cells. Three-fold serial dilutions were performed from 1:12 to 1:8748 and added to 5-250 infectious units of pseudovirus for 1 hour. 50% inhibitory concentration value inverses were used to calculate pNT50 values. This assay has a lower limit of detection (LLD) of 12.

Serum and PBMC samples were analyzed from Healthcare workers (HCWs) following primary series and first monovalent booster doses. For Nursing Home Residents (NHRs), we additionally analyzed samples following second monovalent and bivalent booster doses. We excluded samples collected between a breakthrough infection and the following vaccine dose. Analyzed subjects were classified as infection-naive or prior infection following each dose. At the time of the bivalent dose, we tallied the number of prior vaccines and infections to describe the total prior exposures for each subject.

We identified the intervals between vaccination and sample collection to estimate the decay kinetics of anti-Spike antibodies and neutralizing titers for Wuhan, and BA.4/5 strains. Next, we excluded individuals who developed evidence for a new infection during a given time interval. New or prior infection was determined by history of PCR or antigen test results, presence of existing antibody titers against nucleocapsid protein (NCP), or a rise outside of laboratory variance for anti-spike, receptor binding domain, or neutralizing assay results not accounted for by vaccination history.

We assumed that Day 14 after vaccination showed peak antibody levels and an exponential decay following that peak. Post-vaccine samples were collected on or around day 14 and subsequent samples within 400 days following the same vaccine were analyzed. Given our assumed peak at Day 14, we subtracted 14 from the days between vaccine and sample and imputed zero when this value was negative. Mixed-effects linear models predicting the log-transformed response with days since the peak and a random intercept for each subject were estimated.

For primary series and first monovalent booster doses, Wuhan strain models were estimated with separate decay functions for each dose, subgroup (HCW and NHR), and infection status (naive and prior). Among NHR, additional models were estimated across four doses for each strain, with separate decay functions for each dose and infection status. An exploratory model was considered among NHR following the bivalent booster, comparing decay functions by total number of prior exposures (vaccine or infection) for Wuhan and BA.4/5 strains.

Model estimates were used to calculate half-lives and predicted levels at days 14, 180, and 365 following vaccine dose. Model contrasts were implemented to compare peaks and decay rates across doses and subject subgroups. Unadjusted contrast p-values are presented in supplemental results.

All analysis was performed in R Version 4.2.2 with functions from the nlme and emmeans packages.

## Results

Our NHR cohort maintained a relatively stable older age (median age 74-76 years old) over the series of doses, but with a rising proportion of women (42% female for the primary series and 55% female for 2nd monovalent booster). Women comprised 54% and 59% of the HCW cohort following the primary series and 1st monovalent booster, respectively, with a median age below 50 (Table 1).

**Table 1:**
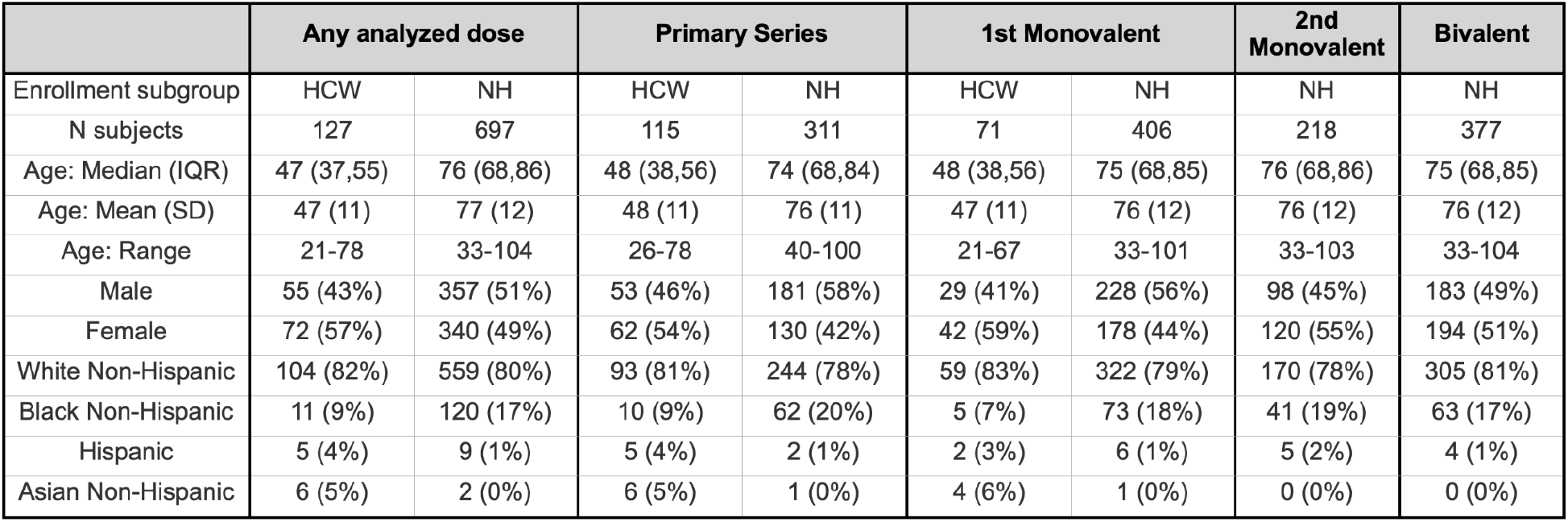
Cohort demographics by vaccine dose.

Among NHRs, we observed a plateau in the model-estimated anti-Spike peak levels across subsequent doses to both Wuhan and Omicron BA.4/5 strains, while estimated peak neutralizing titers increased with each booster dose. NHRs with prior infection show significantly higher peak Wuhan anti-Spike titers than infection-naive NHRs for all doses examined (**Figure 1A)**. However, NHRs with prior infection only show significantly higher model-estimated Wuhan neutralizing titer peaks (**Figure 1B**) than naive NHRs following the primary series (PS) and first monovalent (MV) boost with no difference detected for the subsequent two doses (second MV and first Bivalent (BV)).

**Figure 1:**
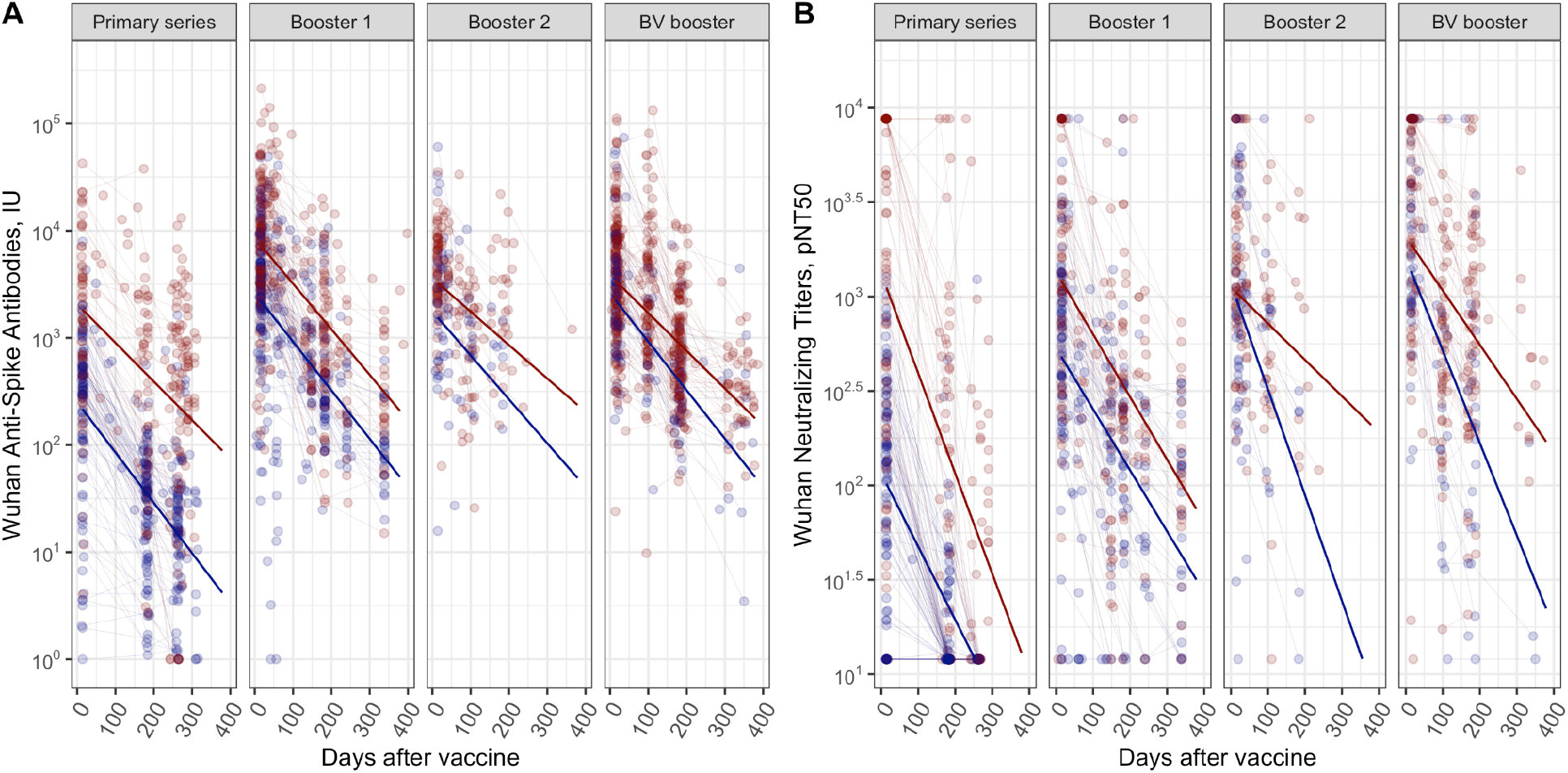
NHR following 4 doses, Wuhan strain for anti-Spike antibodies (A) and neutralizing titers (B). Red dots and lines correspond to subjects with prior infection at the given dose and blue dots and lines correspond to infection-naive subjects.

From these post-vaccine peaks, the model-estimated half-lives of NHR Wuhan anti-Spike titers in both those with and without prior infection across the four examined doses were generally consistent, all falling between 64 to 97 days with longer half-lives predicted for those with prior infection than infection naive for each dose (Table 2). A larger range of half-lives was predicted for Wuhan neutralizing titer decay, where NHRs naive to infection show a less rapid decay following the PS and a more rapid decay following the second MV booster, though with both the upper and lower limits of detection observed for this assay and such truncation impacting estimates. Decay following the second MV booster may also be impacted by the low number of subjects naive to infection at the time of vaccine administration.

**Table 2:** Model-estimated half-lives (days after peak) and titers at days 14, 180, and 365 following vaccine for NHR following 4 doses. Wuhan strain for anti-Spike antibodies and neutralizing titers; 95% confidence intervals presented for all estimated values.

| Assay | status | lastvaxdose | day14 | day180 | day365 | Half-life (days) |
| --- | --- | --- | --- | --- | --- | --- |
| Wuhan Anti-Spike antibodies | Naive | Primary series | 216(162, 288) | 36(29, 44) | 5(3, 7) | 64.1 (56.9, 73.4) |
|  | Prior | Primary series | 1836(1333, 2529) | 462(379, 564) | 99(71, 139) | 83.5 (70.9, 101.5) |
|  | Naive | Booster 1 | 2251(1796, 2822) | 399(317, 502) | 58(38, 90) | 66.5 (58.4, 77.2) |
|  | Prior | Booster 1 | 7428(6012, 9177) | 1463(1196, 1789) | 239(164, 349) | 70.8 (62.6, 81.5) |
|  | Naive | Booster 2 | 1546(1029, 2322) | 322(151, 686) | 56(10, 314) | 73.4 (46.5, 174.1) |
|  | Prior | Booster 2 | 3258(2471, 4296) | 988(656, 1489) | 262(103, 665) | 96.5 (67.8, 167.2) |
|  | Naive | BV booster | 2260(1644, 3106) | 400(287, 556) | 58(31, 107) | 66.4 (55.7, 82.2) |
|  | Prior | BV booster | 3429(2842, 4137) | 891(750, 1058) | 198(143, 275) | 85.4 (74.9, 99.3) |
| Wuhan Neutralizing titer | Naive | Primary series | 102(78, 134) | 23(18, 29) | 4(3, 7) | 77.6 (67.2, 92) |
|  | Prior | Primary series | 1120(827, 1517) | 148(115, 189) | 15(10, 23) | 56.8 (50.4, 64.9) |
|  | Naive | Booster 1 | 481(375, 615) | 140(111, 175) | 35(24, 53) | 93 (78.8, 113.6) |
|  | Prior | Booster 1 | 1217(933, 1587) | 343(273, 430) | 83(56, 124) | 90.8 (76.8, 110.9) |
|  | Naive | Booster 2 | 978(703, 1360) | 115(53, 251) | 11(2, 58) | 53.8 (38.3, 90) |
|  | Prior | Booster 2 | 1054(786, 1413) | 505(299, 852) | 223(69, 717) | 156.4 (85.4, 930.3) |
|  | Naive | BV booster | 1363(952, 1951) | 210(142, 310) | 26(12, 59) | 61.5 (49.6, 80.9) |
|  | Prior | BV booster | 1895(1470, 2442) | 634(498, 806) | 187(111, 315) | 105.1 (82.2, 145.7) |

Despite variations in decay rates, the half-lives of all assays, strains, and populations assessed exceeded 50 days. Model-predicted titers at days 180 and 365 following vaccination can be compared to benchmark levels to describe sustained antibody levels. We considered the NHR infection naive post-PS antibody level as such a benchmark. The model-estimated Wuhan anti-Spike antibodies vaccine level of 216 BAU/mL was consistently observed at day 180 following subsequent doses. Similarly, the post-primary series infection-naive Wuhan neutralizing titer vaccine level was 102 pNT50. This level was exceeded at day 180 but not consistently present at day 365 for subsequent doses (**Table 2**).

Regardless of prior infection status, similar BA.4/5 anti-spike peaks were achieved following the first, second, and BV boosters (Figure 2 and Table 3). NHRs with prior infection achieved higher peak BA.4/5 neutralizing titers than infection-naive NHR following the first MV booster, second MV booster, and BV booster. The BA.4/5 neutralizing titer peaks increased following each dose for both naive and prior infected NHRs (Figure 2B, Table 3). We found typically longer half-lives for those with prior infection for BA.4/5 anti-spike levels and neutralization titers. Notable are the much shorter half-lives of BA.4/5 neutralization titers compared to anti-spike levels after the BV booster with both naive and prior infected individuals.

**Table 3:** Model-estimated half-lives (days after peak) and titers at days 14, 180, and 365 following vaccine for NHR following 3 booster doses. Omicron BA.4/5 strain for anti-Spike antibodies and neutralizing titers; 95% confidence intervals presented for all estimated values.

| Assay | status | lastvaxdose | day14 | day180 | day365 | Half-life (days) |
| --- | --- | --- | --- | --- | --- | --- |
| BA.4/5<br>Anti-Spike<br>antibodies | Naive | Booster 1 | 1082(692, 1691) | 285(212, 383) | 64(40, 103) | 86.2 (69, 114.8) |
|  | Prior | Booster 1 | 1557(1004, 2415) | 822(633, 1069) | 404(274, 596) | 180.3 (121.9, 346.3) |
|  | Naive | Booster 2 | 1366(1025, 1821) | 330(186, 583) | 68(19, 234) | 80.9 (55.9, 146.3) |
|  | Prior | Booster 2 | 2431(1979, 2985) | 1147(845, 1557) | 497(254, 970) | 153.2 (104, 291) |
|  | Naive | BV booster | 962(748, 1237) | 387(298, 503) | 140(91, 217) | 126.4 (101.6, 167.3) |
|  | Prior | BV booster | 1503(1298, 1741) | 964(838, 1110) | 588(463, 747) | 259.4 (200.8, 366.1) |
| BA.4/5<br>Neutralizing<br>titer | Naive | Booster 1 | 74(52, 105) | 35(25, 48) | 15(9, 24) | 154.4 (115.7, 232.1) |
|  | Prior | Booster 1 | 347(245, 490) | 164(122, 221) | 71(46, 112) | 153.9 (115.4, 230.9) |
|  | Naive | Booster 2 | 242(167, 353) | 30(13, 68) | 3(1, 17) | 55.2 (38.9, 94.9) |
|  | Prior | Booster 2 | 730(528, 1010) | 136(77, 240) | 21(6, 73) | 68.6 (49.6, 111.1) |
|  | Naive | BV booster | 706(471, 1058) | 95(61, 147) | 10(4, 24) | 57.4 (46.6, 74.5) |
|  | Prior | BV booster | 1703(1273, 2279) | 369(275, 495) | 67(37, 124) | 75.3 (61.4, 97.4) |

**Figure 2:**
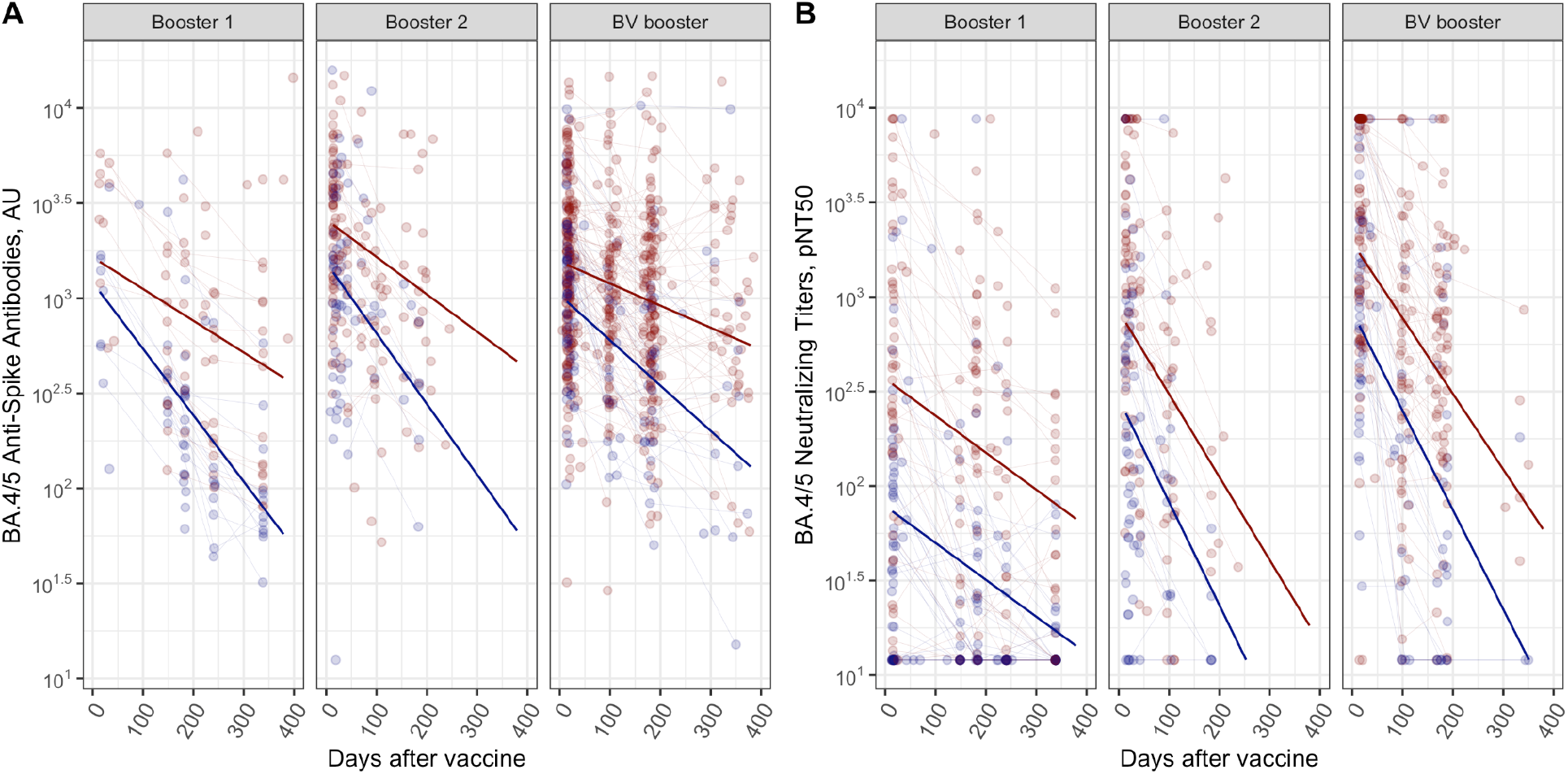
Omicron BA.4/5 in NHR following 3 booster doses for anti-Spike antibodies (A) and neutralizing titers (B). Red dots and lines correspond to subjects with prior infection at the given dose and blue dots and lines correspond to infection-naive subjects.

We constructed a variable that counted the number prior exposures to either the vaccine or infection (Figure 3). The number of exposures ranged from 3 to 6 and were modeled as a categorical variable. In the model results, there was a trend towards higher peak and/or slower decay with each added prior exposure. This was more pronounced in neutralizing titers than in anti-Spike antibodies, but present in both. Subjects with multiple prior infections and 4 prior vaccine doses had the longest-sustained BA.4/5 titers.

**Figure 3:**
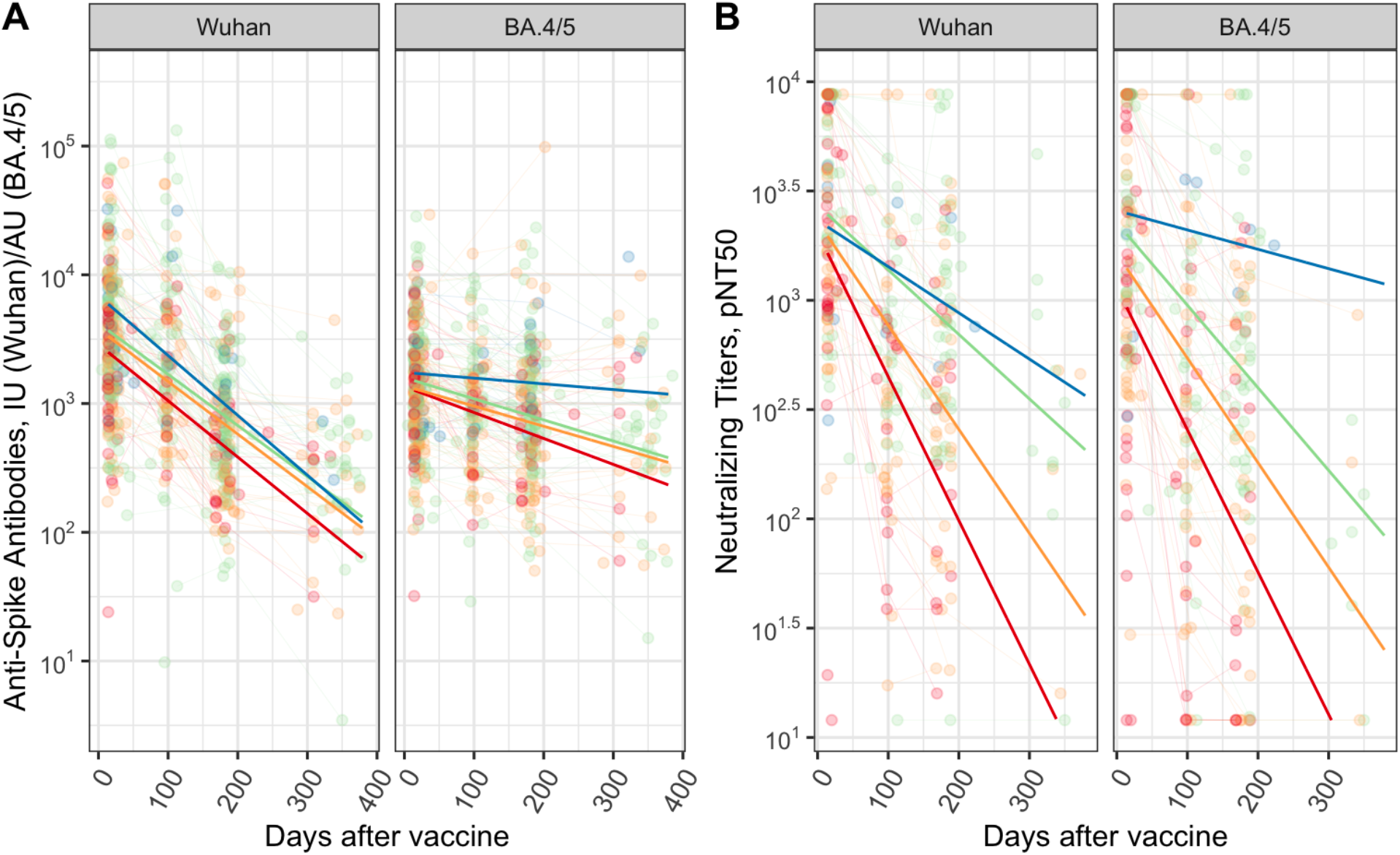
NHR post-Bivalent booster, different peak + decay estimated by # of exposures of vaccine OR infection. Anti-Spike (A) and Neutralizing titers (B) for Wuhan (left), and BA.4/5 (right) following bivalent booster. Number of exposures (sum of infections and prior vaccine doses) ranging from 3 (red), 4 (yellow), 5 (green) and 6 (blue).

We compared whether NHR were different in antibody half-lives following vaccination compared to HCW. The HCW population had a median age of 48 years compared to 75 in NHR. We contrasted the kinetics of decay of antibody levels over time between NHR and HCW in the initial primary vaccine series and first booster since sufficient HCW samples were available. We found a comparable range of half-lives between the NHR and HCW cohorts (**Table 4**). Note vaccination induced much greater antibody levels in NHR with prior infection compared to infection-naive individuals relative to HCWs. Antibody levels after vaccination for infection-naive NHR residents lagged HCW and NHR with prior infection, but then decayed at a similar rate (**Figure 4**). Thus, if a protective antibody level exists, infection-naive NHR would have a much lower likelihood of achieving and sustaining such a post-vaccine protective antibody level compared to HCW and the general population.

**Table 4:** Model-estimated half-lives (days after peak) and titers at days 14, 180, and 365 following vaccine for HCW and NHR following 2 booster doses. Wuhan strain for anti-Spike antibodies and neutralizing titers; 95% confidence intervals presented for all estimated values. Note: NHR values here are also presented in Table 2, repeated here for comparison to HCW.

| Assay | Cohort | Status | Prior dose | day14 | day180 | day365 | Half life (days) |
| --- | --- | --- | --- | --- | --- | --- | --- |
| Anti-Spike antibodies | HCW | Naive | Primary series | 708(507, 990) | 126(94, 169) | 18(11, 31) | 66.7 (56.9, 80.6) |
|  | NHR | Naive | Primary series | 216(162, 288) | 36(29, 44) | 5(3, 7) | 64.1 (56.9, 73.4) |
|  | HCW | Prior | Primary series | 1324(909, 1928) | 210(151, 290) | 27(15, 48) | 62.4 (52.8, 76.4) |
|  | NHR | Prior | Primary series | 1836(1333, 2529) | 462(379, 564) | 99(71, 139) | 83.5 (70.9, 101.5) |
|  | HCW | Naive | Booster 1 | 3097(1739, 5515) | 439(297, 649) | 50(22, 112) | 58.9 (46, 81.6) |
|  | NHR | Naive | Booster 1 | 2251(1796, 2822) | 399(317, 502) | 58(38, 90) | 66.5 (58.4, 77.2) |
|  | HCW | Prior | Booster 1 | 3271(1851, 5780) | 683(462, 1010) | 119(54, 266) | 73.5 (54.8, 111.6) |
|  | NHR | Prior | Booster 1 | 7428(6012, 9177) | 1463(1196, 1789) | 239(164, 349) | 70.8 (62.6, 81.5) |
| Neutralizing titers | HCW | Naive | Primary series | 456(335, 621) | 38(29, 51) | 2(1, 4) | 46.5 (41.3, 53.3) |
|  | NHR | Naive | Primary series | 102(78, 134) | 23(18, 29) | 4(3, 7) | 77.6 (67.2, 92) |
|  | HCW | Prior | Primary series | 963(672, 1382) | 78(57, 107) | 5(3, 9) | 45.8 (40.3, 53.2) |
|  | NHR | Prior | Primary series | 1120(827, 1517) | 148(115, 189) | 15(10, 23) | 56.8 (50.4, 64.9) |
|  | HCW | Naive | Booster 1 | 699(373, 1310) | 197(134, 291) | 48(20, 119) | 91 (60.8, 180.6) |
|  | NHR | Naive | Booster 1 | 481(375, 615) | 140(111, 175) | 35(24, 53) | 93 (78.8, 113.6) |
|  | HCW | Prior | Booster 1 | 697(363, 1335) | 321(220, 470) | 136(59, 312) | 148.8 (83.8, 663.9) |
|  | NHR | Prior | Booster 1 | 1217(933, 1587) | 343(273, 430) | 83(56, 124) | 90.8 (76.8, 110.9) |

**Figure 4:**
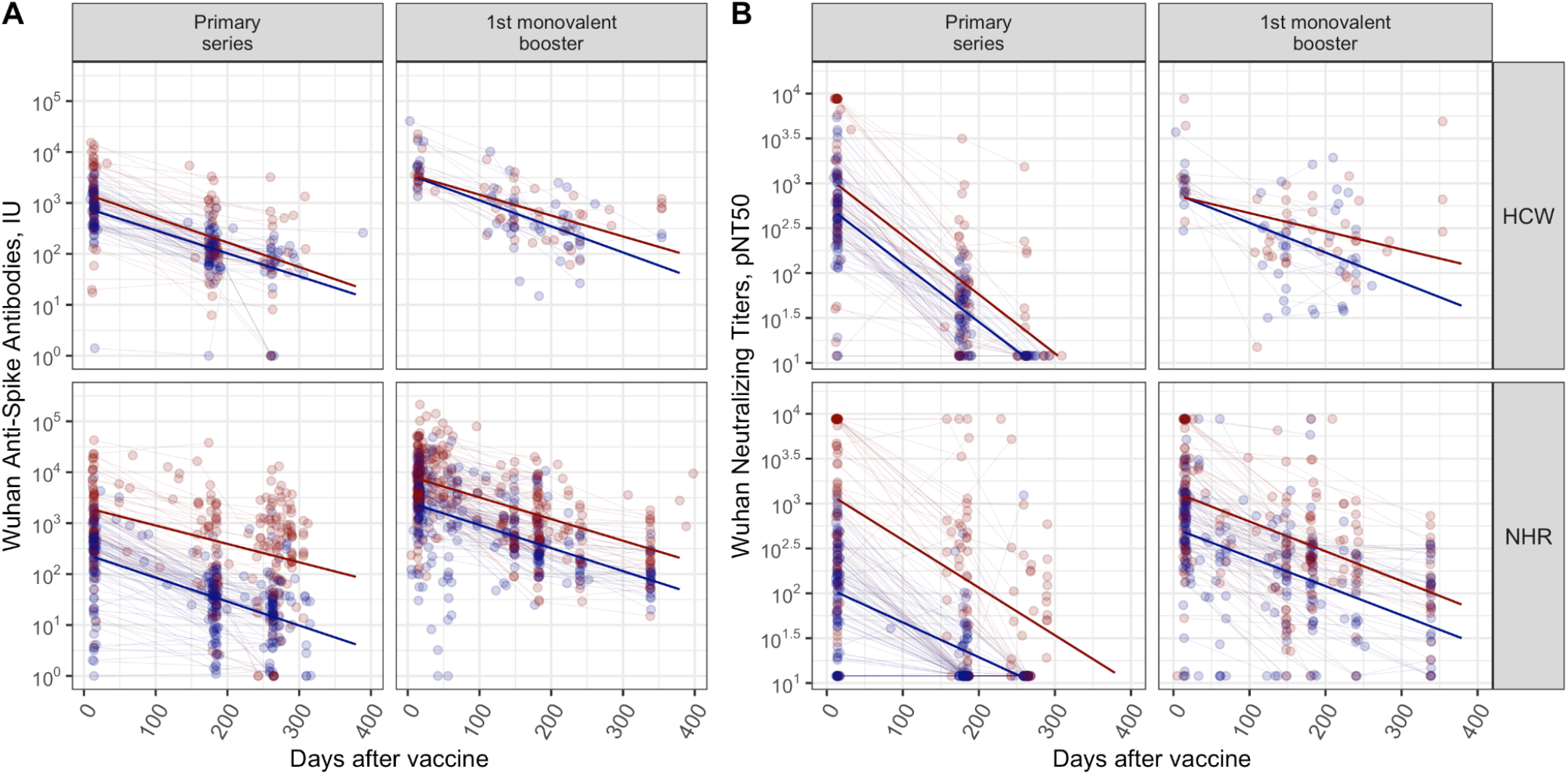
HCW vs. NHR, Wuhan primary series and Booster Wuhan strain for anti-Spike antibodies (A) and neutralizing titers (B). Red dots and lines correspond to subjects with prior infection at the given dose and blue dots and lines correspond to infection naive subjects.

## Discussion

### Impact of Multiple Exposures on Antibody Half-Life

Our findings indicate that multiple exposures to the immune system, whether through vaccination or natural infection, consistently increases antibody half-lives. This likely results from a greater generation of long-lived plasma cells.

Our study also considers the cumulative effect of prior exposures, whether through infection or vaccination, on the long-term trajectory of antibody decay. Examining subjects including the XBB1.5 dose, subjects with multiple prior exposures exhibited higher peak levels and slower decay rates in neutralizing titers compared to those with fewer prior exposures. Chisty et *al*. saw a similar effect of multiple exposures in an earlier study through the first booster dose [15]. Multiple exposures, thus multiple instances of antigenic presentations, likely contribute to the development and refinement of the memory immune response [16-18]. Those that have received more COVID-19 boosters are associated with lower rates of COVID-19 infections [19].

Interestingly, Hofstee et al. [20] highlighted differences in antibody kinetics between infection-naïve and previously infected NH residents. Infection-naïve individuals showed a steeper increase in antibody levels after the third and fifth booster doses compared to those previously infected. However, the antibody levels in previously infected individuals plateaued, possibly due to a negative feedback mechanism or immune imprinting. The faster waning of Omicron-specific antibodies compared to Wuhan-specific antibodies was also noted, indicating the need for ongoing booster vaccinations to maintain protection against emerging variants. Our data also showed a shorter half-life in Omicron-specific titers but this was limited to the neutralization activity.

### Correlation Between Antibody Half-Lives and Vaccine Effectiveness (VE)

We show that antibody half-lives generally correlate with vaccine effectiveness (VE), as discussed in relation to the CDC report. Clinical effectiveness data presented by Dr. Ruth Link-Gelles from the CDC demonstrated a significant reduction in protection from severe disease by day 180, especially in those over age 65 [21, 22]. This supports the current recommendation for a booster 6 months after the prior dose [23]. Notably, data presented by Link Gelles also suggests that loss of protection against infection to levels below 40% occurs more rapidly in the era of Omicron variant strains than earlier in the pandemic. Our immunogenicity data mirror the more rapid decay of the BA.4/5 clinical effectiveness in the neutralization titers after the BV booster, possibly attributable to viral drift [24]. These findings may provide some immunologic mechanisms for these clinical observations.

### Shorter Antibody Half-Lives for Recent Omicron Variants

Our data indicate shorter antibody half-life for more recent Omicron variants than for earlier strains, like for BA.4/5 neutralization titers after the bivalent booster. This decline mirrors that of effectiveness, suggesting that neutralization titers may better serve as immune correlates of protection than binding assay levels [25, 26]. Interestingly, we found the half-life of anti-Spike antibody titers to BA.4/5 after the bivalent booster exceeds that of the BA.4/5 neutralizing titers. Thus, our data also support the idea that neutralization titers provide a better immune correlate of protection than binding assay levels.

### Implications for Vaccination Schedules in NHRs

We interpret these results to support biannual COVID-10 vaccinations for NHRs for now. The observed antibody titer decline beyond day 180 post-vaccination means we need to continue to monitor antibody levels to generate data to justify ongoing biannual booster schedules to maintain protective antibody levels. However, over time, we hope to achieve sufficiently elevated neutralizing levels to justify only annual updates instead. Similarly, Vikström et al. [27] found in their study that the third mRNA vaccine dose elicited a substantial 99-fold increase in S-binding IgG levels in infection-naïve NHR, which corresponded to an increase in neutralizing antibodies. The fourth dose further boosted these levels by 3.8-fold. Despite these increases, the half-life of S-binding IgG was relatively short at 72 days. Importantly, while the vaccine-induced antibodies did not protect against infection with Omicron variants, they were inversely correlated with the risk of death.

### Differences in Immune Response Between NHR and HCW

Our immunologic data from early in the pandemic showed a larger difference in the enhancing effect of prior infection in the NHR group compared to HCW. As the pandemic progressed, the vast majority of individuals, including NHR, had prior infections [10, 28, 29]. The current ACIP booster recommendation of twice yearly SARS-CoV-2 for older individuals is based on increases in severe clinical outcomes in older individuals evident even as early as day 120 post-vaccination, more prominent past day 180. Modeled antibody titer levels in both HCW and NHR show significant declines by day 180 and even more so at day 365, prompting the recommendation of an extra 6-month booster for older individuals.

### Study Limitations

Limitations of this study should be addressed when interpreting these results. This study focuses on antibody responses, while cellular immune responses and their impact on long-term antibody durability were not assessed in this paper. Previous studies have investigated the senescent adaptive immune response in relation to vaccine response, in which T cells that show signs of aging correlate with reduced levels after vaccination and thus may result in a lack of protection against severe COVID-19 [30]. In previous T cell-focused studies, we found that T cell response increases with vaccination doses in this population, but this conclusion was made following the PS, first booster, and second booster [31].

Cohort recruitment took place across nursing homes with a range of resident populations, local outbreaks, and varied vaccine schedules. We did not attempt to address these facility differences, their impact on the population available across doses, or facility-specific vaccine response. This should be taken into account when examining antibody decay between the first boost to the bivalent boost and sex differences.

Our observational design limits our ability to control factors such as variations in vaccination timing and comorbidities. Additionally, the relatively smaller sample size in HCW analyses limits generalizability to broader populations.

## Conclusion

We expect that eventually the combination of serial antigen exposures through vaccinations and infections will result in a sufficiently slower rate of decay in neutralizing antibody levels and higher titers well beyond 6 months. This, coupled with broader subheterotypic immunity, may offer sufficient protection to support recommendations for longer intervals than 6 months between vaccine updates for at-risk populations like NHRs. However, current clinical effectiveness data, our immunologic data, and poor uptake of updated vaccines do not yet support lengthening time between vaccines.

Since COVID-19 vaccine uptake even in the older population is far short of the CDC goals, many individuals are not getting even yearly boosting. Further research on antibody responses should investigate responses beyond one year, particularly in vulnerable populations such as older adults. Studies on how to increase vaccine acceptance are needed as well.

## Data Availability

All data produced in the present study are available upon reasonable request to the authors

## Supplement

Supplemental table: unadjusted p-values of model contrasts comparing peaks and decay rates between infection groups and doses, by cohort and assay

| Cohort | Assay | Dose/infection subset | Contrast | P-value (unadjusted), peak | P-value (unadjusted), decay rate |
| --- | --- | --- | --- | --- | --- |
| NH | Wuhan anti-Spike | Primary Series | Naive - Prior | <.0001 | 0.0149 |
|  |  | Booster 1 | Naive - Prior | <.0001 | 0.523 |
|  |  | Booster 2 | Naive - Prior | 0.0028 | 0.4784 |
|  |  | BV booster | Naive - Prior | 0.0253 | 0.0489 |
|  |  | Naïve | Booster 1 - Primary series | <.0001 | 0.7007 |
|  |  |  | Booster 1 - Booster 2 | 0.09 | 0.7368 |
|  |  |  | Booster 2 - Primary series | <.0001 | 0.6328 |
|  |  |  | Booster 1 - BV booster | 0.9829 | 0.9901 |
|  |  |  | Booster 2 - BV booster | 0.1154 | 0.7428 |
|  |  |  | BV booster - Primary series | <.0001 | 0.7616 |
|  |  | Prior | Booster 1 - Primary series | <.0001 | 0.1471 |
|  |  |  | Booster 1 - Booster 2 | <.0001 | 0.1243 |
|  |  |  | Booster 2 - Primary series | 0.0057 | 0.5166 |
|  |  |  | Booster 1 - BV booster | <.0001 | 0.0572 |
|  |  |  | Booster 2 - BV booster | 0.7372 | 0.5735 |
|  |  |  | BV booster - Primary series | 0.0005 | 0.8446 |
|  | Wuhan neutralizing titers | Primary Series | Naive - Prior | <.0001 | 0.0019 |
|  |  | Booster 1 | Naive - Prior | <.0001 | 0.8517 |
|  |  | Booster 2 | Naive - Prior | 0.7347 | 0.0092 |
|  |  | BV booster | Naive - Prior | 0.1352 | 0.0051 |
|  |  | Naïve | Booster 1 - Primary series | <.0001 | 0.1351 |
|  |  |  | Booster 1 - Booster 2 | 0.0001 | 0.0473 |
|  |  |  | Booster 2 - Primary series | <.0001 | 0.1478 |
|  |  |  | Booster 1 - BV booster | <.0001 | 0.0135 |
|  |  |  | Booster 2 - BV booster | 0.1317 | 0.5788 |
|  |  |  | BV booster - Primary series | <.0001 | 0.1307 |
|  |  | Prior | Booster 1 - Primary series | 0.6402 | <.0001 |
|  |  |  | Booster 1 - Booster 2 | 0.4143 | 0.1118 |
|  |  |  | Booster 2 - Primary series | 0.763 | 0.0002 |
|  |  |  | Booster 1 - BV booster | 0.0068 | 0.3801 |
|  |  |  | Booster 2 - BV booster | 0.0006 | 0.2963 |
|  |  |  | BV booster - Primary series | 0.0045 | <.0001 |
|  | BA.4/5 anti-Spike | Booster 1 | Naive - Prior | 0.2522 | 0.0026 |
|  |  | Booster 2 | Naive - Prior | 0.0013 | 0.0708 |
|  |  | BV booster | Naive - Prior | 0.0025 | 0.0004 |
|  |  | Naïve | Booster 1 - Booster 2 | 0.358 | 0.8118 |
|  |  |  | Booster 1 - BV booster | 0.6136 | 0.0375 |
|  |  |  | Booster 2 - BV booster | 0.0297 | 0.1441 |
|  |  | Prior | Booster 1 - Booster 2 | 0.0583 | 0.635 |
|  |  |  | Booster 1 - BV booster | 0.874 | 0.2504 |
|  |  |  | Booster 2 - BV booster | <.0001 | 0.1126 |
|  | BA.4/5 neutralizing titers | Booster 1 | Naive - Prior | <.0001 | 0.9889 |
|  |  | Booster 2 | Naive - Prior | <.0001 | 0.4591 |
|  |  | BV booster | Naive - Prior | 0.0005 | 0.1048 |
|  |  | Naïve | Booster 1 - Booster 2 | <.0001 | 0.004 |
|  |  |  | Booster 1 - BV booster | <.0001 | <.0001 |
|  |  |  | Booster 2 - BV booster | <.0001 | 0.8704 |
|  |  | Prior | Booster 1 - Booster 2 | 0.0002 | 0.0085 |
|  |  |  | Booster 1 - BV booster | <.0001 | 0.0004 |
|  |  |  | Booster 2 - BV booster | <.0001 | 0.6827 |
| HCW | Wuhan anti-Spike | Primary Series | Naive - Prior | 0.0541 | 0.671 |
|  |  | Booster 1 | Naive - Prior | 0.7766 | 0.5693 |
|  |  | Naïve | Booster 1 - Primary series | <.0001 | 0.5644 |
|  |  | Prior | Booster 1 - Primary series | 0.0039 | 0.6745 |

